# The use of artificial intelligence in the diagnosis of carious lesions: Systematic review and meta-analysis

**DOI:** 10.1101/2024.05.03.24306821

**Authors:** Vanessa Gallego Arias Pecorari, Laís Renata Almeida Cezário, Caio Vieira de Barros Arato, Tainá de Lima Costa, Karine Laura Cortellazzi, Roberto Fiório Pecorari, José Erasmo Silva

**Author notes:** These authors contributed equally to this work. These authors also contributed equally to this work.

## Abstract

**Background:** The use of Artificial Intelligence (AI) has many applications in the healthcare field. Dental caries is a disease with a prevalence rate of over 50% in Brazil. The diagnosis of caries is usually based on a clinical examination and supplementary tests such as X-rays. The accuracy of a diagnostic test is evaluated by its sensitivity, specificity, and accuracy. Various algorithms and neural network configurations are being used for caries diagnosis.

**Objective:** This systematic review evaluated the sensitivity, specificity, and accuracy of using deep machine learning through a convolutional neural network in diagnosing dental caries.

**Methods:** This systematic review was conducted in accordance with the Preferred Reporting Items for Systematic review and Meta-Analyses (PRISMA) 2020 guidelines and registered with Prospero (ID CRD42024411477). We used the PubMed, MEDLINE, and LILACS databases and MeSH and DECs descriptors in the search.

**Results:** After analyzing the eligibility of the articles, we selected 33 for full-text reading and included 13 in the meta-analysis. We used the sensitivity, specificity, accuracy data, and the number of positive and negative tests to generate a 2x2 table with TP, FP, FN, TN rates, and accuracy. We evaluated the heterogeneity of the SROC curve using the Zhou & Dendurkuri I 2 approach. The results showed that the sensitivity and specificity of the machine learning for detecting dental caries were 0.79 and 0.87, respectively, and the AUC of the SROC curve was 0.885.

**Conclusion:** The literature presented a variety of convolutional neural networks [CNN] architecture, image acquisition methods, and training volumes, which could lead to heterogeneity. However, the accuracy of using artificial intelligence for caries diagnosis was high, making it an essential tool for dentistry.

## Introduction

Dental caries is one of the most prevalent diseases (about 50%) in children worldwide and is considered a public health problem (Bagramian, 2009). If not treated in time, it can affect not only chewing function but also speech, smile and the psychosocial environment and quality of life of the child and family (Losso, 2009). Dental caries is a multifactorial disease involving bacterial, dietary and host determinants influenced by multiple sociological and environmental factors (Uribe, 2009; Leong *et al*., 2013).

Diagnosis of the disease Caries is a complex process which involves the interpretation of a set of data from clinical signs and symptoms and complementary exams (Nyvad, 2004; Pretty, 2006). The method for detecting carious lesions must have some characteristics essential to be considered adequate. Be reliable, non-invasive, capable of detecting caries lesions at an early stage and capable of differentiating lesions reversible from irreversible. In addition to affordable cost, comfort for the patient, speed and ease of execution, it must also be viable for all surfaces of the teeth with adequate accuracy (Marinho, 1998).

The radiographic detection of tooth decay is fundamentally based on the fact that with the progression of a tooth decay lesion, the mineral content of tooth enamel and dentin decreases, resulting in an attenuation of the X-ray beams when they pass through the tooth. These characteristics can be observed in the image with increased radiographic density (Silva, 2008). The radiographic examination has high sensitivity in detecting dentin caries lesions but low sensitivity for detecting enamel caries lesions (Soares et al., 2012). The most recommended technique for the radiographic detection of caries is “bitewing,” also known as interproximal (Braga, 2010; Wenzel, 2004). The interproximal radiography allows a better estimation of the more sensitive depth of proximal and occlusal caries in dentin than clinical inspection alone (Purger, 2011)

The artificial neural network [ANN] is the basis of machine learning [“machine learning - ML”] and deep learning [“deep learning - DL”]. A *neural network* is an algorithm composed of layers connected that processes input data, which were extracted from the X-ray image, using convulsive neural networks, and through an activation function, it will classify patients as sick or not sick in the output layer. Artificial intelligence [AI], specifically deep learning using convolutional neural networks [CNNs], has been suggested to improve the reliability and validity of diagnosis by image analysis. CNNs allow mapping an input (image) to an output (classification) based on a set of weights and learned data (LeCun *et al*., 2015). Accuracy above 90% enables the development of software and applications that will assist the dentist in decision-making, enabling the diagnosis of the disease for the population in censuses, public services and private practices.

This work aims to evaluate the sensitivity, specificity and accuracy of using deep machine learning through CNNs in diagnosing dental caries in periapical radiographs of adults through a systematic review.

### Material and methods

Our study protocol was registered on PROSPERO (CRD42024411477). This systematic review and meta-analysis followed the Preferred Reporting Items for Systematic Reviews and Meta-Analyses (PRISMA) standards. Therefore, prior to the beginning of the research, a search was carried out in the literature to find out whether there was a need for a systematic review or whether other reviews that addressed the proposed topic already existed. As we did not find matching work, we drew the protocol for the Systematic registration, and the project followed the criteria recommended by PRISMA and the Minimum information about the clinical artificial intelligence modelling checklist (MI-CLAIM).

### Search strategy

Two authors (VGAP and RFP) separately performed PubMed, MEDLINE, LILACS and Cochrane Library database search using standard search formulas. To search for gray literature (unpublished studies), we used the following sources: ISI Web of Knowledge, British Library Inside, BMC Meeting Abstracts, OpenSIGLE, Clinical Trials database and REBEC. Additionally, we researched the thesis database, government publications and CAPES.

We created the search strategy using subject descriptors (index) for MeSH (Medical Subject Headings) and DECs (Health Science Descriptors). To search for works, we selected terms, the increase in uncontrolled vocabulary with synonymous words, acronyms, related terms and spelling variations (“entry terms”), as shown in tables 1 to 3 in the different bases used. The search rule and Syntax were through Boolean logic that uses logical operators OR, AND and NOT, as shown below:

**Table 1.** Characteristics of selected articles

| Author/ Year | Positive gold standard image (test bench) | Total image (Test bench) | Tooth type | Image acquisition method | Image area | Total images | Total training | Training with cavities | Learning type | Number of layers | Batches | Seasons | Momentum | Learning rate |
| --- | --- | --- | --- | --- | --- | --- | --- | --- | --- | --- | --- | --- | --- | --- |
| Lee et al. 2023 | 300 | 600 | Molar and/or premolar | Periapical X-Ray | Tooth | 3000 | 2400 | 1200 | Transferring learning | 22 | 32 | 1000 | NI | 0.01 |
| Askar et al. 2021 | 374 | 2781 | Incisors and canines | Pictures | ROI | 2781 | . | 374 | . | 8 | 150 | . | 0.9 | 0.00006 |
| Mao et al. 2021 | 350 | 700 | Molar and/or premolar | Bitewing X-Ray | Tooth labeled | 3716 | 700 | 350 | Transferring learning | 25 | 64 | 100 | 0.9 | 0.00006 |
| Cantu et al. 2023 | 128 | 141 | Molar and/or premolar | Bitewing X-Ray | Tooth Labeled | 3686 | 3293 | . | Adaptive moment estimation | NI | 2 | 190 | NI | 0.00005 |
| Schwendicke et al. 2023 | 92 | 226 | Extracted molar and/or premolar | Intraoral Picture | Tooth | 226 | 226 | 92 | Transferring learning | 50 | NI | NI | NI | 0.00001 |
| Hur et al. 2021 | 322 | 792 | Molar | Panoramic X-Ray and tomography | Tooth | 2642 | 2400 | NI | Transferring learning | NI | NI | NI | NI | 0.0001 |
| Duong et al. 2021 | 514 | 587 | Molar and/or premolar | Smartphones Pictures | Tooth | 587 | NI | 293 | Transferring learning | 50 | NI | 10 | NI | 0.0001 |
| Vinayahalingam et al. 2021 | 50 | 100 | Third molar | Panoramic X-Ray | Tooth | 500 | 320 | 200 | Transferring learning | 18 | 32 | 10 | NI | 0.0001 |
| Li et al. 2022 | 200 | 300 | All types | Periapical X-Ray | ROI | 4129 | 3829 | 2062 | Cross-entropy | 18 | 50 | 200 | NI | 0.0001 |
| Kühnisch et al. 2022 | 83 | 479 | All types | Nikon Pictures | Tooth | 2417 | 1891 | 348 | Transferring learningn / cross-entropy | 18 | 16 | 50 | NI | 0.001 |
| Estai et al. 2022 | 329 | 862 | Molar and/or premolar | Bitewing X-Ray | ROI | 8618 | 7756 | 2967 | Transferring learning | 50 | 64 | 10 | NI | 0.001 |
| Park et al. 2022 | 150 | 300 | Molar and/or premolar | Intraoral Picture | ROI | 1938 | 1638 | 678 | Cross-entropy | 18 | NI | 20 | NI | 0.001 |
| Kim et al. 2022 | 300 | 2000 | Molar and/or premolar | Panoramic X-Ray | ROI | 10 | NI | NI | Softmax | NI | NI | 10 | NI | 0.001 |

ROI regions of interest. NI not informed.

### VHL

MH:“Dental **Caries” OR (Tooth Decay) OR** (Tooth Decay) **OR (** Cavities **) OR (** Dental Cavities **) OR (** Dental Cavities **) OR (** Decayed Tooth **) OR (** Carious Lesions **) OR (** White Spots **) OR (Dental Caries) OR (** Caries **) OR (** Caries Dentales **) OR (** Carious Lesions **) OR (** White Spots Dentales **) OR (Dental Caries) OR (** Caries, Dental **) OR (** Carious Dentin **) OR (** Carious Dentins **) OR (** Carious Injury **) OR (** Carious Injuries **) OR (** Decay, Dental **) OR (** Dental Decay **) OR (** Dental White Spot **) OR (** Dental White Spots **) OR (** Dentin, Carious **) OR (** Dentins, Carious **) OR (** Lesion, Carious **) OR (** Lesions, Carious **) OR (** Spot, Dental White **) OR (** Spots, Dental White **) OR (** White Spot, Dental **) OR (** White Spots, Dental **) AND MH:” Machine Learning” OR (Machine Learning) OR (** Machine Learning **) OR (** Transfer Learning **) OR (** Machine Learning **) OR (** Machine Learning **) OR (** Transfer Learning **) OR (Automatic Learning) OR (** Transfer Learning ) _ **OR ( Machine Learning) OR (** Learning, Machine **) OR (** Learning, Transfer **) OR (** Transfer Learning )

### PUBMED

“Dental Caries”[Mesh] OR **(Dental Caries) OR (** Caries, Dental **) OR (** Carious Dentin **) OR (** Carious Dentins **) OR (** Carious Lesion **) OR (** Carious Lesions **) OR (** Decay, Dental **) OR (** Dental Decay **) OR (** Dental White Spot **) OR (** Dental White Spots **) OR (** Dentin, Carious **) OR (** Dentins, Carious **) OR (** Lesion, Carious **) OR (** Lesions, Carious **) OR (** Spot, Dental White **) OR (** Spots, Dental White **) OR (** White Spot, Dental **) OR (** White Spots, Dental ) AND “Machine Learning”[Mesh] OR **(Machine Learning) OR (** Learning, Machine **) OR (** Learning, Transfer **) OR (** Transfer Learning )

### Eligibility Criteria

We included the literature published in PubMed, MEDLINE, LILACS, and the Cochrane Library database and gray literature on diagnosing dental caries in adults using AI in radiographic images.

The inclusion criteria was as follows: Participants: patients with occlusal and/or interproximal dental caries of both sexes in permanent dentition who were diagnosed using interproximal or periapical X-ray imaging; Index test: studies that used the use of artificial intelligence as an index test; Reference test: Visual diagnosis by specialists in the field; Outcomes: sensitivity, specificity and accuracy and studies that evaluated caries in percentage or qualitatively classifying in presence and absence; Types of studies: observational cross-sectional studies, cohort, case-control and clinical trials.

Excluded criteria were case reports and animal studies, decayed primary teeth, studies that analyzed artificial intelligence for the diagnosis of diseases other than dental caries in the field of dentistry knowledge, studies that the response variable (outcome) or parameters have not been clearly reported, impossibility of extracting study data and lack of response after contact with the study authors.

### Study selection

Before searching for articles, we calibrated reviewers to ensure high reliability in selecting works. The references of the selected articles were listed using the Rayyan reference manager <http://rayyan.qcri.org/reviews/5 >. We removed the duplicate articles after adding the search results from all databases. Two reviewers (VGAP and RFP) selected the articles, analyzing them by titles and abstracts and reached a consensus on the disagreements between reviewers. We created a clinical form containing the eligibility criteria to record the reasons for exclusion from each study using the flowchart. We excluded ineligible studies based on the eligibility criteria. After selecting the works, we obtained the full articles.

### Data extraction

The following data were extracted from the included literature: reference with the first Author and year of publication, type of study, classification of study quality (risk of bias), number of images used in articles, type of neural network used (architecture, number of neurons), hyperparameters adopted, dental caries was categorized in presence or absence, sensitivity of the diagnostic method, specificity of the diagnostic method, accuracy of the method or area on the ROC curve.

### Quality of articles

Two independent reviewers (VGAP and RFP) classified the articles by the instrument of critical evaluation and their risk of bias using the tool Quadas 2 (“Quality Assessment of Diagnostic Accuracy Studies-2”). Reviewers rated each work by consensus based on this tool to qualify the studies.

### Statistical analysis

We used the sensitivity and specificity data, with the number of positive tests obtained by the “gold” standard and the number of tests to generate a 2x2 table with the TP, FP, FN and TN rates. With these rates, we recalculated the sensitivity and specificity values with their respective 95% confidence intervals and presented them in forest graphs (Forest Plots). As it is common that the presence of heterogeneity in diagnostic accuracy studies is not only by chance but also by the implicit variation of the cutoff point (threshold effect), the summary point that represents the sensitivity and rate of false positives (1-specificity) were combined using a bivariate model of random effects (Reitsma et al., 2005). We chose the bivariate model of random effects because it considers the correlation between the rates of sensitivity and false positives of the included studies. We evaluated the heterogeneity of the summarized ROC curve [SROC] by the I2 approach of Zhou & Dendurkuri, implemented in the Mada package. In addition to the summary point of sensitivity and rate of false positives (1-specificity), we also calculated the Positive Likelihood Ratio and Negative (RV+ and RV-) together with the estimate of the Diagnostic Odds Ratio [DOR] through the MCMC procedure of Zwindermann et al. (2008) parameters obtained by the bivariate model. We performed all analyses in R language (version 4.2 for Mac iOS) with the help of Mada (version 0.5.11) and DTAplots (version 1.0.2.5) packages.

### Quality assessment and publication bias

To evaluate the quality of the studies, we applied the QUADAS-2 risk checklist to test the bias risk in each study (Table 2). The articles selected presented a low risk of bias. Most of the authors followed the STARD and/or CLAIM standards. QUADAS2 presents four domains (patient selection, Index test, reference test and follow-up over time).

**Table 2.** Risk of bias of selected studies

| Author | Year | Domain 1 |  | Domain 2 |  | Domain 3 |  | Domain 4 | Bias Risk |
| --- | --- | --- | --- | --- | --- | --- | --- | --- | --- |
|  |  | A | B | A | B | A | B | A |  |
| Lee et al. | 2023 | LOW | LOW | LOW | LOW | LOW | LOW | LOW | LOW |
| Askar et al. | 2021 | LOW | LOW | LOW | LOW | LOW | LOW | LOW | LOW |
| Mao et al. | 2021 | LOW | LOW | LOW | LOW | LOW | LOW | LOW | LOW |
| Cantu et al. | 2023 | LOW | LOW | LOW | LOW | LOW | LOW | LOW | LOW |
| Schwendicke et al. | 2023 | LOW | LOW | LOW | LOW | LOW | LOW | LOW | LOW |
| Hur et al. | 2021 | LOW | LOW | LOW | LOW | LOW | LOW | LOW | LOW |
| Duong et al. | 2021 | LOW | LOW | LOW | LOW | LOW | LOW | LOW | LOW |
| Vinayahalingam et al. | 2021 | LOW | LOW | LOW | LOW | LOW | LOW | LOW | LOW |
| Li et al. | 2022 | LOW | LOW | LOW | LOW | LOW | LOW | LOW | LOW |
| Kühnisch et al. | 2022 | LOW | LOW | LOW | LOW | LOW | LOW | LOW | LOW |
| Estai et al. | 2022 | LOW | LOW | LOW | LOW | LOW | LOW | LOW | LOW |
| Park et al. | 2022 | LOW | LOW | LOW | LOW | LOW | LOW | LOW | LOW |
| Kim et al. | 2022 | LOW | LOW | Unclear | Unclear | LOW | LOW | LOW | Unclear |

## Results

Based on the search strategy, 225 articles were selected from the databases. Among these, 78 duplicates were removed, and 57 were excluded for other reasons, resulting in 90 articles. Upon reviewing the title and abstract, 57 papers were excluded for not meeting the eligibility criteria, 2 did not receive responses from the authors, leaving 31 for full-text reading. Of these 31, 13 articles were deemed eligible for meta-analysis (Figure 1).

**Fig 1.**
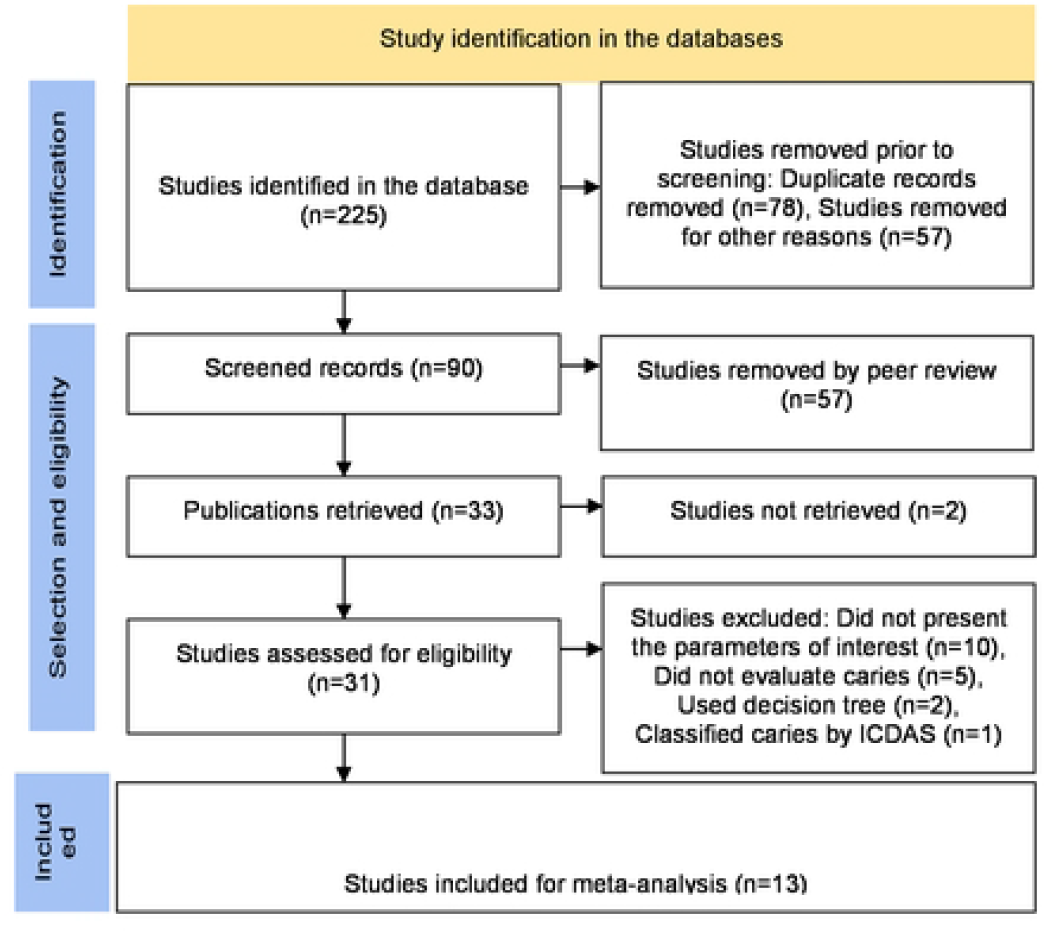
Flowchart.

In total, true positive (TP), false positive (FP), false negative (FN), and true negative (TN) rates from 13 studies were utilized to generate the Forest Plot illustrating sensitivity and specificity. The sensitivity of machine learning for caries detection ranged from 0.58 to 0.90 across studies, and specificity ranged from 0.68 to 0.95. The test for equality was statistically significant for both sensitivity (χ2=188.34; p<0.0001) and specificity (χ2=332.37; p<0.0001) with a weak negative correlation (ρ=−0.071) between sensitivity rates and false positives (1-specificity), as demonstrated in Figure 2.

**Fig 2.**
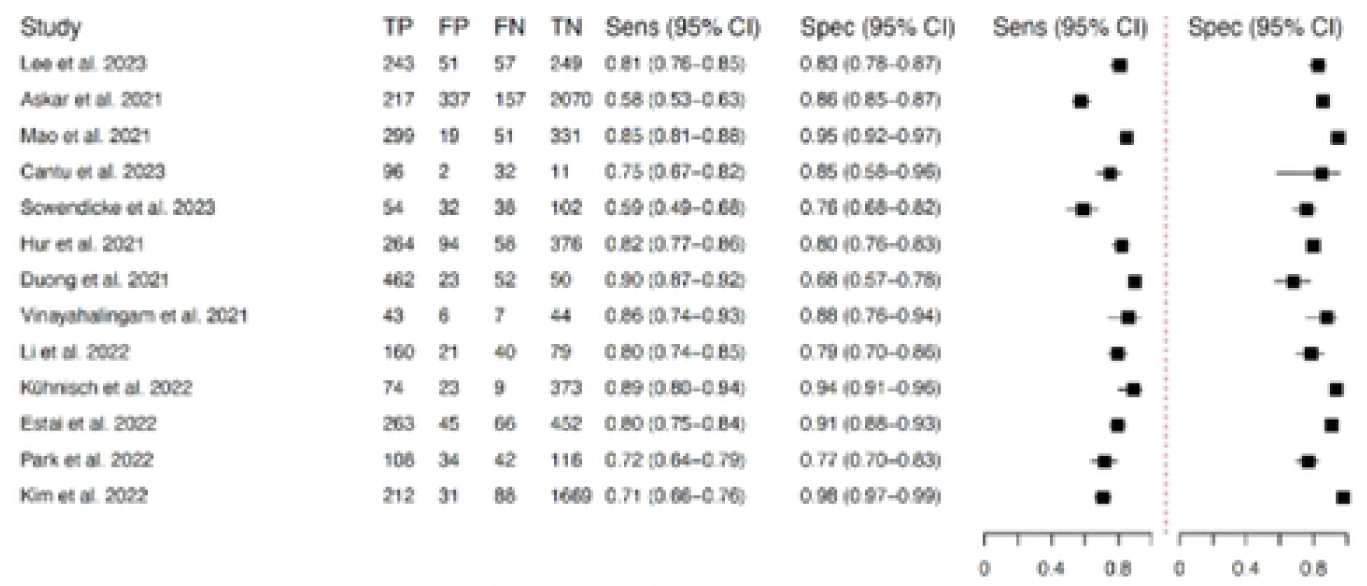
Forest Plot of machine learning accuracy measures in dental caries detection.

The result obtained by the bivariate random-effects model revealed that the sensitivity and specificity of machine learning for detecting dental caries were 0.79 (95% CI: 0.73; 0.84) and 0.87 (95% CI: 0.81; 0.92), respectively. The summary receiver operating characteristic (SROC) curve’s area under the curve (AUC) was 0.885, and the heterogeneity, estimated by the I2 statistic using Zhou & Dendukuri’s approach, was I2 = 45.3% (Figure 2).

Figure 3 depicts the SROC, with the summary point of sensitivity and false positive rate (1-specificity) of machine learning for detecting dental caries. The SROC demonstrated an accuracy of 0.885 encompassing all studies within the prediction region. We calculated the Positive and Negative Likelihood Ratios along with the Diagnostic Odds Ratio estimate as presented in Table 3.

**Table 3.**
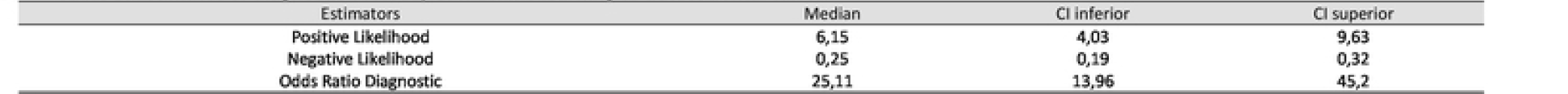
Estimates of diasnostic accuracy of machine learning to detect dental caries.

| Estimators | Median | CI inferior | CI superior |
| --- | --- | --- | --- |
| Positive Likelihood | 6,15 | 4,03 | 9,63 |
| Negative Likelihood | 0,25 | 0,19 | 0,32 |
| Odds Ratio Diagnostic | 25,11 | 13,96 | 45,2 |

**Fig 3.**
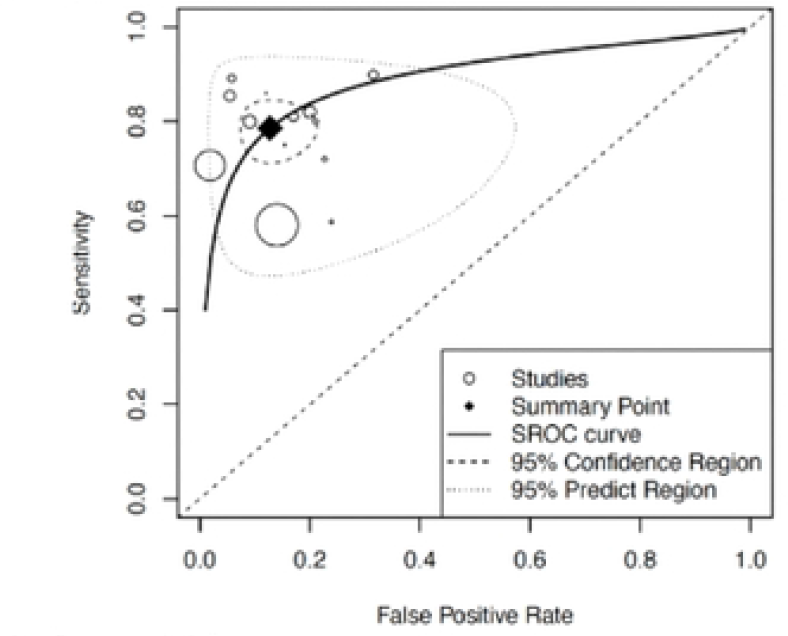
SROC (“Summary Receiver-Operating Characteristic”).

Additionally, based on the data, we constructed the Fagan’s nomogram, as shown in Figure 4. The Fagan’s nomogram illustrated that for a positive caries test diagnosed by convolutional neural networks, the probability of the tooth having caries is 86.01%, and the probability of it being a false negative when caries is not detected is 20%.

**Fig 4.**
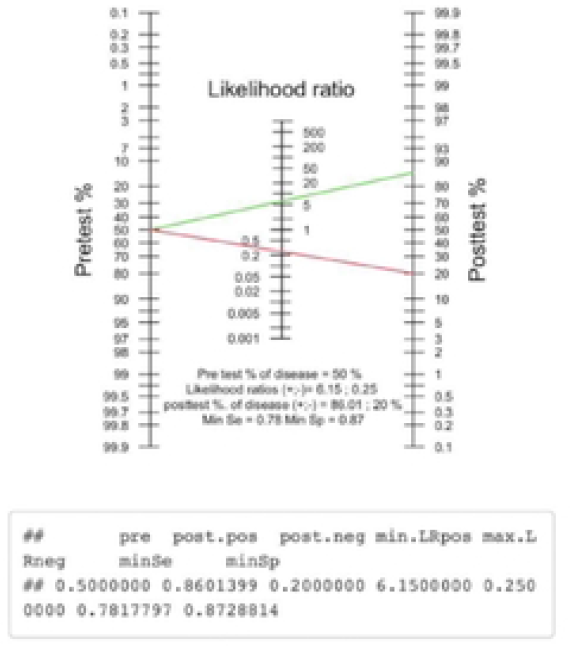
Fagan’s nomogram

## Discussion

Among the machine learning techniques until 2018, the supervised vector method [SVM] was the most used classification method. According to Santana et al. (2022) SVM have classification algorithms with better sensitivity values when compared to ANN. However, SVM and CNNs result in similar conclusions. CNNs are a class of deep learning algorithms that have been widely used in computer vision tasks, including caries detection. One of the main advantages of CNNs is the ability to detect relevant characteristics at different spatial scales, which is especially important for detecting caries, such as in X-ray images or smartphone photos.

SROC demonstrated an accuracy of 0.885, containing all studies within the prediction region and a summarized sensitivity of 0.79 (95%CI 0.73 to 0.84) and summarized specificity of 0.87 (95%CI 0.81 to 0.92) and heterogeneity of 45.3%. Of the articles selected for the meta-analysis, five authors worked with regions of interest [ROIs] to reduce dimensionality and make the diagnosis more accurate. However, sensitivity ranged from 0.58 to 0.80 and specificity from 0.77 to 0.98, not the only way to improve accuracy.

The sensitivity results among the authors ranged from 0.68 to 0.95. The two authors with the lowest sensitivity values were Askar et al. (2021) and Scwendicke et al. (2023), and the authors with the highest sensitivity values were Kühnisch et al. (2022) and Duong et al. (2021). The authors who presented the lowest specificity were Duong et al. (2021) and Park et al. (2022), and the highest specificity values were the works of Estai et al. (2022) and Kim et al. (2022). There is an inverse relationship between sensitivity and specificity. Our review observed this relationship with the authors Duong et al. (2021) and Askar et al. (2021).

The clinical validity of caries diagnosis is complex because it requires a clinical diagnosis in which it needs to present clean, dry and well-lit teeth. Rx is considered a complementary examination to the clinical examination and requires experience from the dentist. Thus, even being considered the gold standard is not without error. The X-ray quality, as contrast and proper positioning, is another preponderant factor for a correct diagnosis. In all articles, dentists diagnosed from photos or X-rays, making the diagnosis less accurate in more subtle cases, such as white spots and caries without cavities, regardless of the evaluator’s expertise, which can also impact machine learning.

The type of image in our study consisted of both photos of teeth of smartphones and images of X-ray (panoramic or interproximal). They are different images because the photos of smartphones and high-resolution cameras are coloured, while those from X-ray have grayscale. The authors who used camera photos were Askar et al. (2021), Schwendicke et al. (2023), Duong et al. (2021), Kühnisch et al. (2022) and Park et al. (2022). The other authors used X-ray (bitewing or panoramic) or CT scans. This diversity of images may be one of the explanations for the heterogeneity among the authors. The hyperparameters the authors used showed similarities in the learning rate (between 0.001 and 0.00001). The number of times varied considerably among the authors (10 to 10,000). There were also variations between the number of batches (2 to 150).

Most authors used Transfer Learning and cross-entropy as a method of deep learning. Another important factor to highlight and may be related to the differences in sensitivities and specificity of AIs learning is the network architecture adopted in each study. The high number of layers with fewer images can cause overfitting. Of the selected studies, 23.08% (3 articles) used 50 layers, 30.77% (4 articles) used 18 layers, and 15.38% (2 articles) used 22-25 layers of artificial neurons. The author who presented high sensitivity and specificity values was Kühnisch et al. (2022), who used 1891 images of all types of teeth in training the network architecture of 18 layers and the type of learning of Transfer Learning and cross-entropy.

The sample size is a crucial point for machine learning. The sample size and proportion of the disease are parameters that alter diagnostic test sensitivity and specificity values (Linnet et al. 2012). The volume of images in training is one of the main factors for adequate learning and, consequently, better accuracy values. However, having a database with large volume, veracity, and image quality is the most significant difficulty encountered in studies. There were large variations between authors regarding the number of images obtained and augmented for training. The images were enlarged by delimiting regions of interest, rotating among other techniques, and may have improved sensitivity and/or specificity. However, new studies with different severities of caries disease need to be carried out to improve the accuracy of CNN, especially in initial caries where the diagnosis is difficult.

The articles selected presented a low risk of bias since most of the authors followed the STARD and/or CLAIM standards. QUADAS2 presents four domains (patient selection, Index test, reference test and follow-up over time). In the case of artificial intelligence use, several domains receive low risk because, for the first domain, a large number of patients and images are obtained from several sources randomly. As for the index and pattern domains, all images pass the pattern for the index test to learn to diagnose. Thus, a low risk of bias was attributed. However, it is worth remembering that QUADAS2 is a tool to assess the risk of bias from conventional diagnostic tests and is not being developed for diagnostic test analysis for AIs. With the increasing use of IA in diagnostic tests, there is a need to develop a specific tool for these cases in systematic reviews. Only the author Kim et al. (2022) classified it as uncertain due to the incomplete description of the network architecture of the index test and the network training method. However, it is worth remembering that the tool still needs to be specific to evaluate articles that use artificial intelligence as an index test.

The DOR measures the effectiveness of the diagnostic test analyzed and is independent of the disease’s prevalence, unlike the test’s accuracy. The AIs presented a DOR of 25.11 times of the test when giving positive results of the individual with the disease concerning those without the disease and a 95% confidence interval ranging from 13.96 to 45.20. We can also verify through the Fagan nomogram that the CNNs presented the probability of 86.1% of diagnosing existing caries and only 20% of false negatives.

Despite the values obtained in our study, the improvement of the sensitivity, specificity and accuracy of CNNs can still be explored in future studies with the impact analysis of the number of training images, types of images, prevalence of disease and different architectures in the diagnosis of caries, enabling its use in modern dentistry with greater safety.

## Conclusion

Using artificial intelligence through convolutional neural networks to diagnose caries showed global accuracy (0.88), and it can be trained to detect visual patterns that indicate the presence of caries in both X-ray images and photos and can be used in new devices to assist the dentist in diagnosing caries disease.

## Data Availability

PubMed, MEDLINE, LILACS, and the Cochrane Library database

https://pubmed.ncbi.nlm.nih.gov/

https://bvsms.saude.gov.br/minibanners/medline/

https://lilacs.bvsalud.org/

https://www.cochranelibrary.com/

